# A Core Outcome Set for Small Abdominal Aortic Aneurysms

**DOI:** 10.1101/2024.12.06.24318626

**Authors:** Sabrina L.M. Zwetsloot, Lotte Rijken, Igor Koncar, Marina Dias-Neto, Regent Lee, Riikka Tulamo, Christian-Alexander Behrendt, Fabien Lareyre, Noeska N. Smit, Stefan P.M. Smorenburg, Venkat Ayyalasomayajula, Vincent Jongkind, Kak Khee Yeung, VASCUL-AID collaborators & expert consensus meeting collaborators

## Abstract

**Background:** It remains elusive which patients with an abdominal aortic aneurysm (AAA) will experience cardiovascular disease progression. Current literature on small AAAs is characterized by heterogeneous and selective outcome reporting. A core outcome set (COS), a consensus-based list of outcomes that should be reported as a minimum in research on a specific disease, is therefore needed. This is especially important for research using artificial intelligence (AI) techniques. The current study aimed to create a European COS for use in clinical and AI research on small AAAs. This COS will be used in the VASCUL-AID project, which aims to identify risk factors for cardiovascular disease progression in patients with small AAA using AI.

**Methods:** This COS was developed in line with Core Outcome Measures in Effectiveness Trials (COMET) initiative recommendations in three steps. First, a longlist of AAA outcomes was identified through a systematic literature search and through focus groups with patients, their caregivers, and healthcare professionals. Second, a three-round European Delphi survey was conducted with patients and a wide range of healthcare professionals. Third, an expert consensus meeting with key opinion leaders (KOL), patients, and patient representatives was held to finalize the COS.

**Results:** The AAA longlist consisted of 91 outcomes; 73 outcomes identified from the systematic literature search and 18 patient-centred outcomes identified from the focus groups. A total of 257 participants (of which 104 were patients) participated in the Delphi study. Delphi round 3 was filled in by 221 (66.9%) participants. The highest-rated outcomes from the third Delphi round were included for discussion in the expert consensus meeting, which was attended by 23 KOL, two patients and one patient representative. Ten core outcomes were chosen across six health domains, with AAA rupture, mortali-ty, health-related quality of life, clinical success, and graft infection attaining 100% consensus for inclusion in the COS.

**Conclusion:** This first COS for small AAAs consists of ten outcomes and should be implemented in all clinical and AI research on small AAA.

**Clinical perspective:** *What is new?:*

- Current literature on small abdominal aortic aneurysms (AAA) is characterized by heterogene-ity and selective outcome reporting.
- A Core Outcome Set (COS) is an expert consensus-based list of outcomes that should be re-ported as a minimum in all clinical research on a specific disease.
- A COS for small AAA was created, consisting of ten outcomes across six health domains, which may also be applied in artificial intelligence (AI) research such as the VASCUL-AID project.

*What are the clinical implications?:*

- This COS for patients with small AAA may enhance uniformity across and reproducibility of clinical research.
- In addition to clinical research, this COS may also be applied in cardiovascular risk prediction models for small AAA patients and will be included as endpoints in the VASCUL-AID pre-diction models.

## Introduction

Patients with an abdominal aortic aneurysm (AAA) suffer from a progressively degenerating and dilating abdominal aortic wall that may ultimately rupture. Treatment consists of adequate cardiovascular disease management and surgical repair at a recommended abdominal aortic diameter above 55 mm in men and 50 mm in women.^1^ Risk factors associated with AAA are, amongst others, smoking, prior myocardial infarction, high blood pressure and positive family history for AAA.^2,3^ Individuals show great variation in AAA growth rate and rupture risk, which impedes adequate personalized risk prediction.^4,5^ The great variation in outcome reporting in AAA literature presents another challenge in personalized treatment strategies, thwarting comparison between clinical trials.^6^ Appropriate outcome reporting by use of a pre-defined set of outcomes chosen by clinicians and patients is crucial for research to be reproducible and clinically relevant.

Core outcome sets (COSs) serve an important purpose in enhancing uniformity and reproducibility of research. A COS comprises a list of expert consensus-based outcomes that should be reported in all clinical research on a certain disease.^7^ Patients are intently involved in the process of COS development, because patient and clinician views on the importance of certain outcomes do not always align.^8,9^ Within vascular surgery, COSs have recently been created for outcomes after infrarenal AAA repair and patients undergoing major lower limb amputation, but one for patients with small AAAs (diameter below threshold for surgical repair) under routine surveillance is lacking.^10,11^ The current study aimed to develop a COS for outcomes on small AAAs.

Notably, this COS will be used in the VASCUL-AID project (HORIZON-HLTH-2022-STAYHLTH-01-two-stage); a European consortium that aims to develop artificial intelli-gence (AI) models for prediction of cardiovascular disease progression and rupture risk in patients with a small AAA for personalized treatment strategies.^12^ A defined set of core out-comes is especially important in AI research, as the standardization of variables enables col-lection of higher-quality data from heterogeneous medical data sources.^13^ Therefore, a COS on small AAA is imperative to guide the research within the VASCUL-AID studies and other AI studies on small AAA.

## Methods

### Data availability

The data that support the findings of this study are available from the corresponding author upon reasonable request.

### Study design

This COS has been developed in accordance with the Core Outcome Measures in Effectiveness Trials (COMET) initiative recommendations and according to the COS-STAR principles.^14,15^ Moreover, the COS was pre-registered in the COMET database (on Oct 19th, 2023; study number 2856) and the protocol for this COS development is available online.^16^

The COS was developed in three steps. First, a longlist of AAA outcomes was created. Second, a three-round Delphi study was conducted. Third, expert consensus meetings with key opinion leaders (KOL), patients, and patient representatives were held. An overview of the full study design is shown in Fig. 1. The study process was reviewed throughout by the study management group consisting of SZ and senior authors VJ and KY and adjustments to the study design were made accordingly.

**Figure 1:**
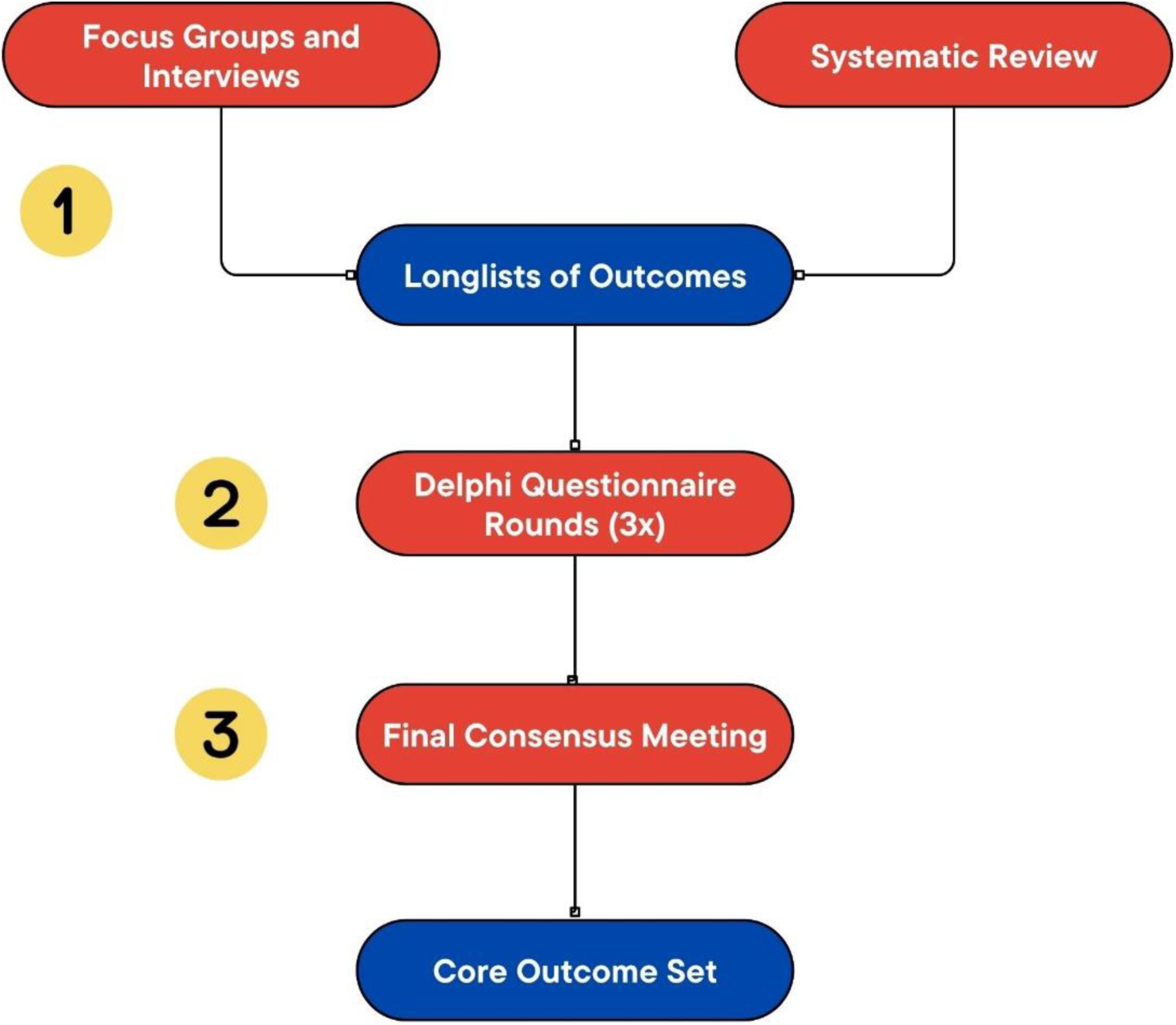
Overview of the study design to create a core outcome set for abdominal aortic aneurysms (AAA). 1) A longlist of AAA outcomes was obtained through a systematic literature review and focus groups with stakeholders (patients and healthcare professionals). 2) A three-round Delphi study was conducted with stakeholders. 3) A final consensus meeting with key opinion leaders and patients was carried out to finalize the definitive core outcome set.

### Ethics

The Medical Research Ethics Committee of the Amsterdam University Medical Centres concluded that the current study is not covered by the scope of the Dutch Medical Research Involving Human Subjects Act (WMO). In the Netherlands, written informed consent on an informed consent form was obtained from all participants of the focus groups and Delphi study. For all other participating countries, informed consent was obtained by phone or e-mail.

### Stakeholders

Stakeholders were recruited from eight European countries from the VASCUL-AID consortium^12^: the Netherlands, United Kingdom, Finland, Norway, Serbia, Germany, Portugal, and France. In every participating centre, a local coordinator (a vascular surgeon) was responsible for participant recruitment. Stakeholders were recruited through purposive sampling to maximize variety of participants. Patients were eligible if they had a diagnosis of AAA with a diameter 30 – 55 mm or if they had previously been treated for AAA. For the focus groups and Delphi study, a wide range of healthcare professionals (HCPs) from the field of vascular surgery and cardiovascular medicine were invited to participate. For the expert consensus meeting, two to four KOL from each participating centre were invited to participate.

### AAA outcome longlist

The AAA outcome longlist was comprised of all unique outcomes identified from a systematic literature review on AAA outcomes and focus groups with stakeholders. A total of 5,611 studies were identified in the systematic literature search, of which 612 full texts were screened and 380 studies were found eligible for inclusion (manuscript submitted). Finally, from the 264 unique outcomes that were identified, 77 were included in the AAA longlist. These outcomes covered 27 domains from the Dodd taxonomy, which is a 38-item taxonomy designed for outcome standardization in medical literature.^17^

Focus groups were conducted to identify additional patient-centred outcomes. They were 60-90 minutes long, followed a semi-structured format with a grounded theory approach, and were moderated by vascular surgery PhD researchers and/or vascular surgeons in three European countries (the Netherlands, United Kingdom and Serbia). The aim was to include between six and ten participants per focus group.^18^ Focus groups were held in participants’ native language. Guiding questions addressed the following topics: treatment success and/or complications, quality of life, and disease progression, including participants’ attitudes towards these topics. Thematic content analysis was done using MaxQDA software (VERBI Software, 2021).^19^ Two researchers independently coded and categorized the transcripts in an analytical framework into themes and subthemes.^11^ Newly-identified unique patient-centred outcomes were added to the AAA longlist.

### Delphi study

Patients from the Netherlands, United Kingdom, Serbia, and Portugal and HCPs from all eight consortium centres participated in the Delphi study. An ethical waiver was obtained in the Netherlands, however, in Germany, Finland, France, and Norway ethical approval from their own ethical committee proved necessary. Therefore, no patients could be recruited form these sites. If invited eligible patients and healthcare professionals filled in the first round of the Delphi, they were considered participants and were included in the study. If, in subsequent rounds, they failed to fill in the Delphi round, they were removed from the study. Patient characteristics were collected by local coordinators at every centre. HCPs provided their own baseline characteristics through a short survey.

Questionnaires were distributed in participants’ native language, with the exception of Norwegian and French HCPs, who received English questionnaires. The questionnaires were written in English and Dutch and were subsequently translated into German, Portuguese, Serbian, and Finnish with AI-based translation software DeepL.^20^ Translations were checked by the local coordinator of each respective centre. A plain language description, reviewed by a layperson unfamiliar with vascular terminology, accompanied each outcome. With regard to dispersion of the surveys, local coordinators were offered three different options: online distribution through Castor Electronic Data Capture,^21^ a web-based data management platform that enables distribution of surveys via e-mail; postal dissemination; and interviews with participants (by phone or through a visit).

The number of Delphi rounds was prespecified as three. Consensus definitions are shown in table 1. Outcomes were presented in the chronological order of the Dodd taxonomy domains.^17^ Participants were able to score an outcome on a 9-point Likert scale from 1 (“unimportant”) to 9 (“extremely important”). In Delphi round 2 and 3, participants were shown the previous round’s median results of every outcome for both stakeholder groups separately. In addition, in round 1 and 2, participants were able to suggest additional outcomes that they felt were missing. The outcomes that did not meet the predefined consensus criteria were removed from and suggested outcomes that were considered new were added to the consecutive Delphi round. After the last Delphi round, the list of twenty outcomes that attained the highest percentage of 7-9 scores was distributed to the expert consensus meeting participants.

**Table 1:** Consensus definitions for in-and exclusion of outcomes shown per Delphi round.

| Round | Consensus definition |
| --- | --- |
| 1 | *Consensus-in:* >50% of respondents in both stakeholder groups scored an outcome 7- |
|  | 9.<br>*Consensus-out*: if >15% of participants scores the outcome as 1-3. |
| 2 | *Consensus-in*: >70% of respondents in both stakeholder groups scored an outcome 7-9.<br>*Consensus-out*: if >15% of participants scores the outcome as 1-3. |
| 3 | The twenty outcomes with the highest percentage of consensus will be discussed in the expert consensus meetings. |

Baseline characteristics were analysed using descriptive statistics, including mean and median scores and range, using Microsoft Excel.^22^ Differences between mean and median scores for stakeholder groups collectively were analysed using SPSS version 28.^23^

### Expert consensus meeting

Two different expert consensus meetings were held: one with patients and patient representa-tives, and one with KOL within the field of vascular surgery and cardiovascular medicine. The aim of the expert consensus meetings was to obtain a six-to-eight outcome list per stake-holder group which would afterwards be combined to form one definitive COS.^24,25^ The ex-pert consensus meetings started with a general, anonymous vote through a web-based voting system on all twenty remaining outcomes from the Delphi study.^26^ Outcomes that attained 100% consensus were included in the COS without discussion. The ten remaining outcomes that attained the most consensus, were discussed among the participants of the two consensus meetings, with the aim of coming to a mutual agreement of which to include in the COS.

## Results

### AAA outcome longlist

The AAA outcome longlist consisted of 77 clinical and patient-centred outcomes identified from the systematic literature review and eighteen patient-centred outcomes identified from the focus groups (see Supplementary Table S1 for the full list of patient-centred outcomes). From the 95-item longlist, three items were removed due to being not AAA-specific (“Trau-ma”, “Dementia”, and “Cancer”), one item (“In-hospital infection”) was removed because more specific infections were already included, and one item was merged with another out-come (“Ventilatory assistance” was filed under “(Acute) respiratory failure”). In addition, “Return to work” was combined with an outcome identified from the focus groups and thus changed to “Being able to do (voluntary) work”. Supplementary Table S2 shows the definitive 91-item AAA longlist.

In total, nineteen patients, eleven caregivers, and eighteen HCPs participated in the focus groups (see Supplementary Table S3 and S4 for number and baseline characteristics of partic-ipants). Notably, the majority of patients were male, and patients’ mean age was considerably lower in Serbia (59.4 years vs. 74 years and 76.2 years in the Netherlands and United King-dom, respectively). Fourteen patients (73.7%) were under preoperative surveillance. The per-centage of male HCPs differed between countries, with the Netherlands and the United King-dom including 33.3%, while Serbia included 83.3%.

### Delphi study

The Delphi study initiated in May 2024 with a total of 99 patients and 149 HCPs participating in the first Delphi round (see Fig. 2 for an overview of the full Delphi study). All HCPs as well as patients from the U.K. and the Netherlands filled in the survey online. A minority of Dutch patients (n=8/27; 29.6%) preferred being sent paper questionnaires. Patient interviews in Serbia and Portugal were carried out by a vascular surgeon, vascular surgery resident, or a medical student. Ten additional participants were introduced in round two, due to informed consent forms being received late in the Netherlands (n=2), and U.K. and French participants being sent the first Delphi survey only in the second week (n=8). HCP participants from round one from Portugal and France were invited to participate again in round three due to low participation rate from these countries, which resulted in three additional participants in round three. The last Delphi round was held in June 2024, with a response rate of 78.7% among patients and 81.7% among HCPs.

**Figure 2:**
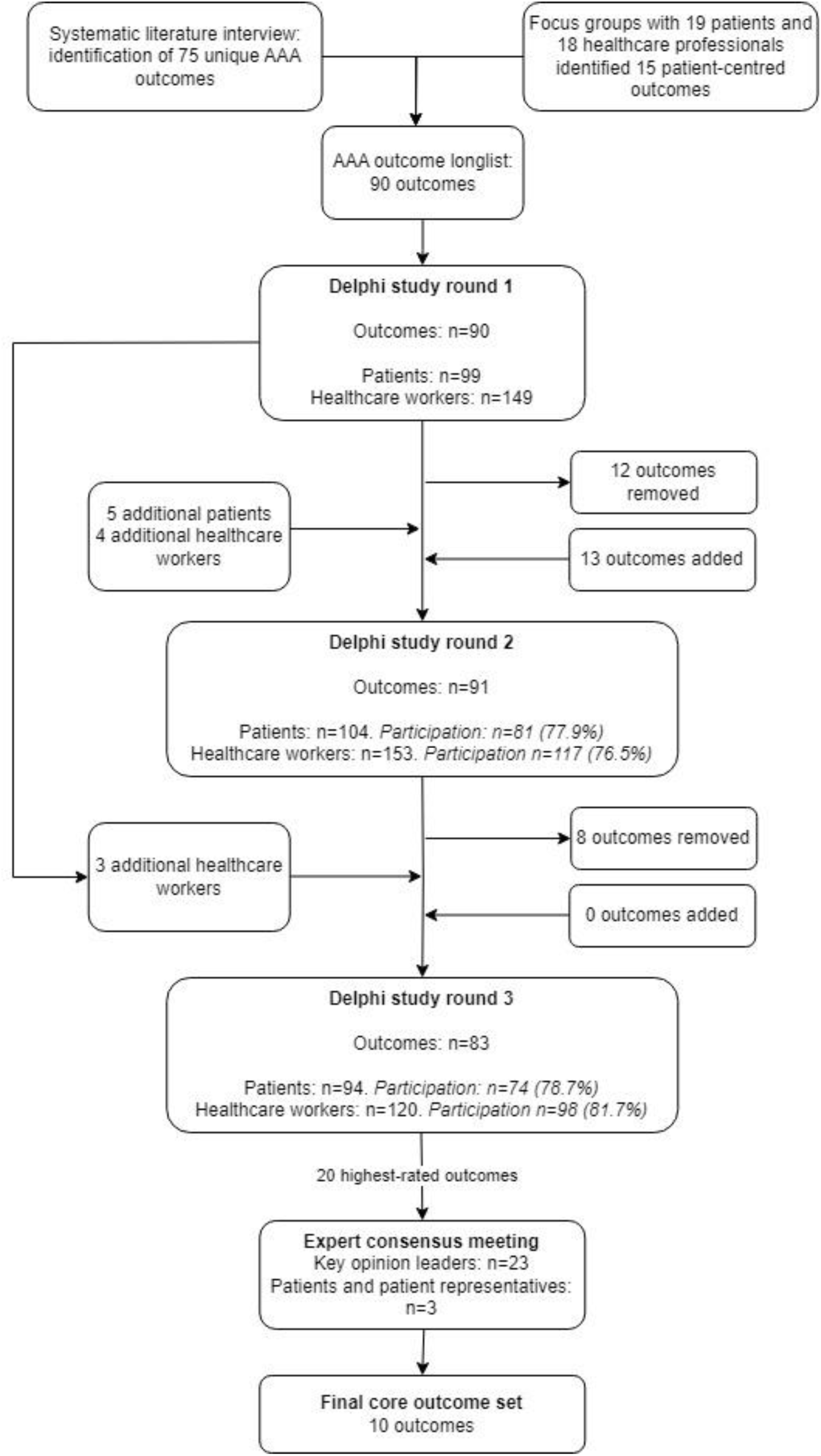
Overview of the full study design with rates of participation and outcomes added and removed per Delphi round. AAA = abdominal aortic aneurysm.

The majority of participating HCPs was a vascular surgeon or vascular resident (n=108, 42.0%), see Table 2 for a full overview of participants’ occupation). The majority of participants were male (64.5% of HCPs and 88.5% of patients). Mean patient age was 72.1 years (SD 6.6). In total, 47 patients (45.2%) were under preoperative routine surveillance, all others had received an intervention.

**Table 2:** Baseline characteristics of participants of the Delphi study. Values are shown as mean (standard deviation) or percentages. NL = the Netherlands, RS = Serbia, PT = Portugal, UK = United Kingdom, FI = Finland, DE = Germany, FR = France, NO = Norway. AAA = abdominal aortic aneurysm.

| Stakeholder | NL | RS | PT | UK | DE | FI | FR | NO | Total |
| --- | --- | --- | --- | --- | --- | --- | --- | --- | --- |
| **Patients** |  |  |  |  |  |  |  |  |  |
| AAA patient | 29 | 39 | 30 | 6 |  |  |  |  | 104 |
| **Healthcare professionals** |  |  |  |  |  |  |  |  |  |
| Vascular surgeon | 13 | 17 | 25 |  | 15 | 7 | 5 |  | 82 |
| Vascular surgery resident | 3 | 18 | 1 | 3 | 1 |  |  |  | 26 |
| Medical doctor in the (vascular) surgery ward | 2 |  |  | 1 |  | 1 | 2 |  | 6 |
| Nurse in the (vascular) surgery ward | 7 | 1 |  | 1 |  | 2 |  |  | 11 |
| Specialised nurse / physician assistant vascular surgery | 2 |  |  | 3 |  |  |  |  | 5 |
| Vascular (diagnostic) technician | 3 |  |  |  |  |  |  |  | 3 |
| Perioperative vascular surgery nurse |  |  | 1 |  |  |  |  |  | 1 |
| (PhD) Researcher vascular surgery | 2 |  |  | 1 |  |  | 1 |  | 4 |
| Medical student |  |  |  |  | 1 |  |  |  | 1 |
| Clinical physiotherapist | 1 |  |  |  |  |  |  |  | 1 |
| Internal medicine specialist | 1 |  |  |  |  |  |  |  | 1 |
| Radiologist |  | 4 |  |  |  |  |  |  | 4 |
| Anaesthesiology resident |  |  |  | 1 |  |  |  |  | 1 |
| Perioperative medicine specialist |  |  |  | 1 |  |  |  |  | 1 |
| Palliative care nurse specialist | 1 |  |  |  |  |  |  |  | 1 |
| Consultant in Medical Biology |  |  |  |  |  |  | 1 |  | 1 |
| Professor in medical imaging |  |  |  |  |  |  |  | 1 | 1 |
| Unknown |  |  |  | 1 | 2 |  |  |  | 3 |
| **Total stakeholder group** | **64** | **79** | **57** | **18** | **19** | **10** | **9** | **1** | **257** |

Table 3 shows the twenty outcomes that attained the highest scores per stakeholder group in the last Delphi round. AAA rupture had the highest mean overall score both by patients and by HCPs (8.93 and 8.80 out of 9.00, respectively). The biggest difference in scores was observed for health-related quality of life (HRQoL), with patients scoring this outcome 1.04 points higher than HCPs. Patients generally awarded higher mean scores to outcomes than HCPs in the last Delphi round (7.85 vs. 7.11 out of 9; p<0.001).

**Table 3:** Overview of the twenty outcomes that attained the highest degree of consensus. Shown are the percentage of participants that awarded a score of 7-9 on each outcome and the mean score given to each outcome. AAA = abdominal aortic aneurysm.

| Outcome | % of scores 7-9 | Mean healthcare worker score | Mean patient score |
| --- | --- | --- | --- |
| AAA rupture | 98.8 | 8.80 | 8.93 |
| Clinical success after an operation | 97.5 | 8.00 | 8.62 |
| Prolongation of lifespan | 97.5 | 8.55 | 8.65 |
| Technical success after an operation | 96.3 | 8.36 | 8.72 |
| Short-term reintervention | 96.3 | 7.76 | 8.61 |
| The AAA can be treated through endovascular treatment | 95.7 | 8.25 | 8.58 |
| The AAA can be treated through open surgical abdominal aortic aneurysm treatment | 95.7 | 8.30 | 8.63 |
| Aortoenteric fistula after an operation | 95.7 | 7.90 | 8.48 |
| Graft infection after an operation | 95.7 | 7.87 | 8.34 |
| Primary patency | 95.7 | 7.75 | 8.52 |
| Good communication by healthcare worker about diagnosis, secondary prevention, and treatment options and complications | 95.7 | 7.87 | 8.48 |
| Cerebrovascular accident, including transient ischemic attack | 95.1 | 7.63 | 8.58 |
| Symptomatic abdominal aortic aneurysm | 95.1 | 7.91 | 8.56 |
| Health-related quality of life | 95.1 | 7.37 | 8.41 |
| Adherence to prescribed drugs | 95.1 | 7.76 | 8.55 |
| Multiple organ failure | 94.6 | 7.84 | 8.54 |
| Neurologic deficit, incl. spinal cord ischemia | 94.5 | 7.80 | 8.35 |
| Amputation | 94.5 | 7.64 | 8.63 |
| Endoleak after endovascular treatment of an AAA | 94.4 | 7.62 | 8.16 |
| Interdisciplinary healthcare worker collaboration | 94.4 | 7.23 | 8.12 |

After the third and last Delphi round, three changes to the outcome list were made: “short-term reintervention” became the more general “re-intervention”; “symptomatic AAA” was changed to “pain from a (symptomatic) AAA” to distinguish between clinical signs of im-pending rupture and actual AAA rupture; and “prolongation of lifespan” was changed to “mortality” to cover both the concept of life extension and mortality.

### Expert consensus meeting

The expert consensus meetings were held on June 10^th^, 2024 in Porto, Portugal. Participants were either present physically or online via Microsoft Teams.^27^ Two moderators per meeting facilitated discussion both online and on the physical location. The spoken language was English. In total, 23 KOL, two patients and one patient representative (from the Dutch Heart Foundation (*Harteraad*)) partook in the expert consensus meetings (Table 4 shows their specifics). Most KOL were from the Netherlands (n=9) and most were vascular surgeons (n=13; 56.5%). After initial voting and discussion, final voting was performed. The outcomes that received at least two-thirds of the votes for “include” were included in the final COS.

**Table 4:**
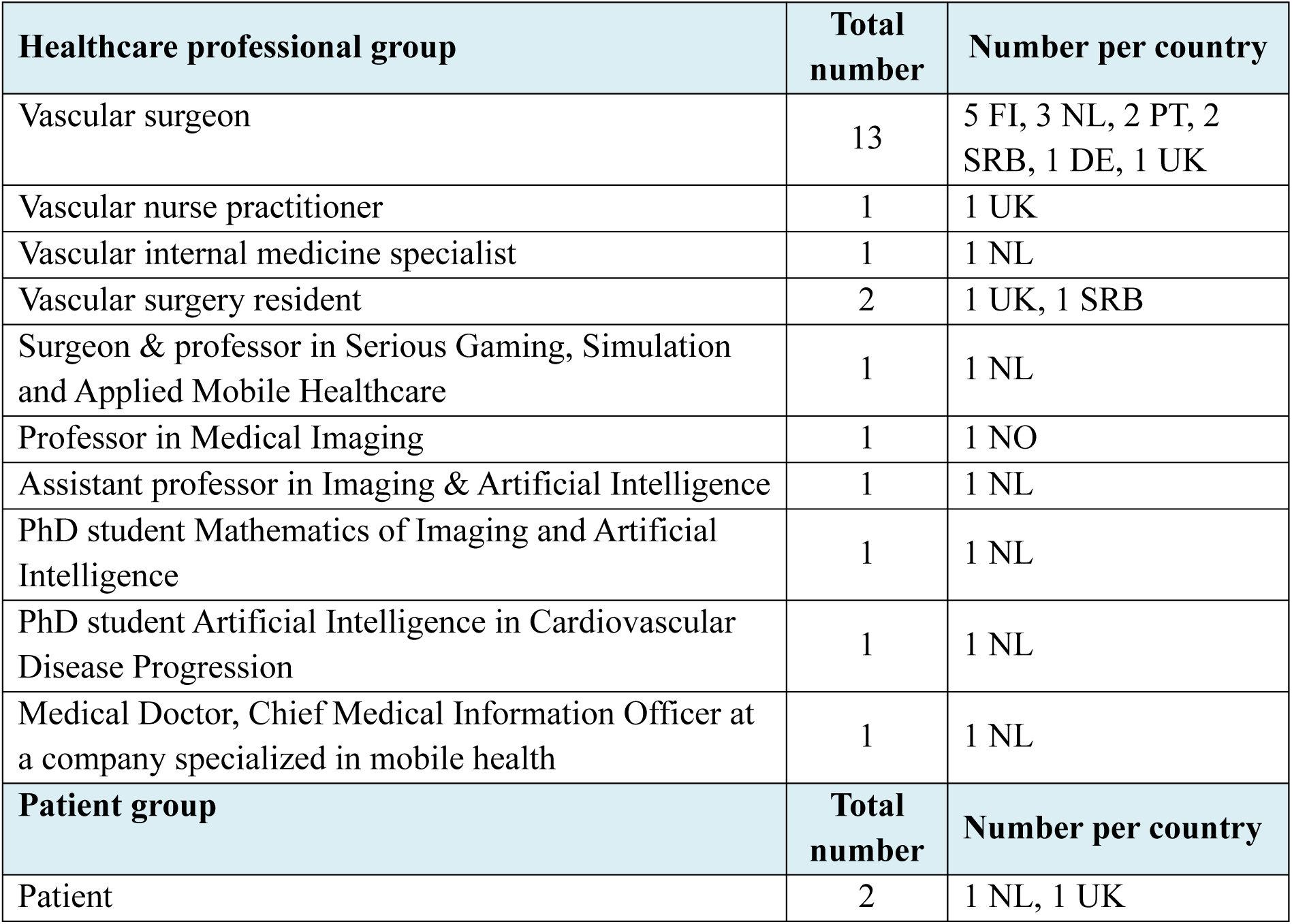
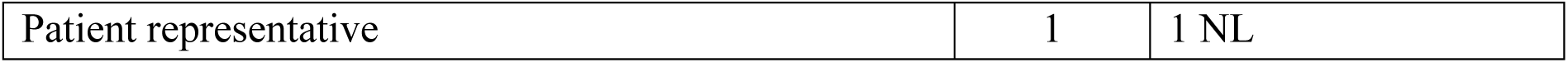
Number and specification of key opinion leaders, patients, and patient representative participating in the expert consensus meeting on abdominal aortic aneurysms. Number of participants per country are shown. FI = Finland, NL = the Netherlands, PT = Portugal, SRB = Serbia, DE = Germany, UK = United Kingdom, NO = Norway.

| Healthcare professional group | Total number | Number per country |
| --- | --- | --- |
| Vascular surgeon | 13 | 5 FI, 3 NL, 2 PT, 2 SRB, 1 DE, 1 UK |
| Vascular nurse practitioner | 1 | 1 UK |
| Vascular internal medicine specialist | 1 | 1 NL |
| Vascular surgery resident | 2 | 1 UK, 1 SRB |
| Surgeon & professor in Serious Gaming, Simulation and Applied Mobile Healthcare | 1 | 1 NL |
| Professor in Medical Imaging | 1 | 1 NO |
| Assistant professor in Imaging & Artificial Intelligence | 1 | 1 NL |
| PhD student Mathematics of Imaging and Artificial Intelligence | 1 | 1 NL |
| PhD student Artificial Intelligence in Cardiovascular Disease Progression | 1 | 1 NL |
| Medical Doctor, Chief Medical Information Officer at a company specialized in mobile health | 1 | 1 NL |
| Patient group | Total number | Number per country |
| Patient | 2 | 1 NL, 1 UK |
| Patient representative | 1 | 1 NL |

### Final core outcome set

Table 5 shows the two stakeholder COS. Four outcomes are common to both COS, namely AAA rupture, mortality, HRQoL, and cerebrovascular accident. The outcomes that attained 100% consensus were AAA rupture, mortality, and HRQoL, clinical success, and graft infection. The final COS consists of ten outcomes across six Dodd domains (see Fig. 3).^17^

**Figure 3:**
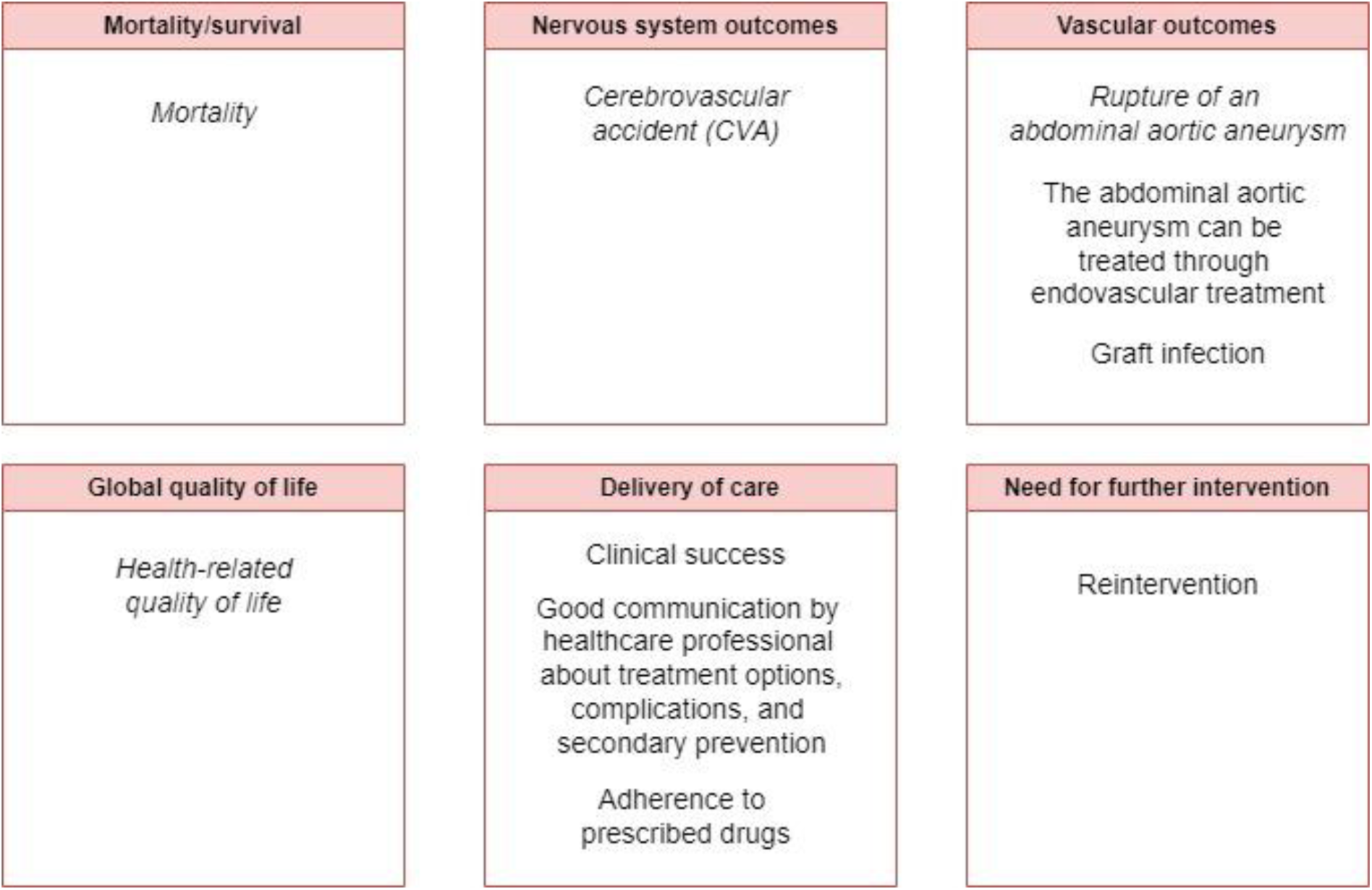
The final core outcome set for small abdominal aortic aneurysms categorized into Dodd taxonomy domains. In italics the outcomes that were included in both the patient and the healthcare stakeholder group.

**Table 5:** The stakeholder (patient and HCP) core outcome sets that were established during the expert consensus meetings in order of most consensus. In bold the outcomes that achieved 100% consensus. In italics the outcomes that are included in both core outcome sets. AAA = abdominal aortic aneurysm.

| Key opinion leader core outcome sets |  |
| --- | --- |
| Healthcare professional | Patient |
| ***AAA rupture*** | ***AAA rupture*** |
| ***Mortality*** | ***Clinical success after an operation*** |
| ***Health-related quality of life*** | ***Graft infection after an operation*** |
| *Cerebrovascular accident* | *Health-related quality of life* |
| Adherence to prescribed drugs | *Mortality* |
| Good communication by healthcare worker about diagnosis, secondary prevention, and treatment options and complications | *Cerebrovascular accident* |
|  | Reintervention |
|  | The AAA can be treated through endovascular treatment |

## Discussion

This is the first COS on small AAAs under preoperative routine surveillance. This European COS consists of ten patient-and healthcare worker-consensus-based outcomes that should be reported as a minimum in all research on small AAAs, the purpose of which is to enhance validity and standardization of literature. Yet, this COS does not restrict the number of outcomes that could be measured and reported in a study.^7^

A strength of this study is the European collaboration and consensus on a large scale, involving 104 AAA patients from four different countries and 153 HCPs from eight different countries. Patient involvement increases the clinical relevance of outcomes.^7^ Thus, the study design of the current study was devised so that patient views were acknowledged; focus groups with patients were conducted, 40% of the Delphi study participants were patients, and separating patients from KOL during the expert consensus meeting ensured that core outcomes chosen by patients were definitely included in the final COS. Moreover, a wide variety of HCPs contributed, ranging from experts in AI to clinical physiotherapists to vascular surgeons.

The current COS is comprised of ten outcomes across six health domains. Two other COSs within vascular surgery have been created: one on outcomes following elective AAA repair and one on outcomes following lower limb amputation.^10,11^ The COS on outcomes following intact AAA repair consists of six outcomes, of which three are also included in the current COS: “Mortality”, “AAA rupture”, and “Quality of life”.^10^ Interestingly, although “Major adverse cardiovascular events” (MACE), a composite outcome denoting acute myocardial infarction, cardiovascular death, and stroke, is often reported in clinical AAA research, its component “myocardial infarction” did not reach enough consensus in the present study.^28^ Likewise, it does not appear in the other two COSs in vascular surgery, which corroborates that this outcome is not considered core.^10,11^ Use of composite outcomes contributes to heterogeneity in research due to varying definitions across research.^28^ Instead, the current COS includes only distinct, individual outcomes.

Differences exist between the patient and KOL core outcomes. Three of the included outcomes in the patient expert meeting are intervention-related outcomes (“Graft infection”, “Clinical success”, and “Reintervention”), all applicable both to open surgical repair and endovascular aortic repair. Untreated AAA patients value complications and re-intervention highly in assessing treatment options.^29^ Likewise, in the third Delphi round, patients awarded a considerably higher mean score to “Short-term reintervention” than HCPs (8.61 vs. 7.76, respectively). In addition, “The AAA can be treated endovascularly” signifies patient preference for endovascular treatment, an attitude that also came forward in our focus groups and is corroborated by literature.^30^ Another point of consideration is the patient-chosen outcome “Graft infection”, a complication associated with high morbidity but relatively low incidence (0.5-5%).^1^ Patients expressed their preference for the inclusion of outcomes they wished to avoid, such as “Graft infection” and “Reintervention”, in the COS.

Outcomes that were chosen exclusively by KOL were “Adherence to prescribed drugs” and “Good communication by healthcare worker”. Although “shared decision-making” (SDM) was also included in the AAA longlist, it did not reach enough consensus. In SDM, the clinician informs the patient on treatment options and complications, resulting in mutual agreement on a treatment decision.^31^ Health literacy in AAA patients is generally low, which may be associated with lower income and lower education.^32,33^ Moreover, AAA patients are often inadequately informed about treatment options.^34^ Inclusion of “Good communication” in the current COS may thus pave the way towards accurate assessment of whether patients feel sufficiently informed by their healthcare provider, as is laid down in law by the concept of “informed consent”.

It is imperative that uptake of this COS is stimulated in clinical research on small AAA. Current literature is heterogeneous and lacks systematic outcome reporting.^6,35,36^ Consistent reporting of core outcomes could decrease outcome reporting bias and application of the same endpoints facilitates evidence synthesis.^37,38^ Lastly, expert-and patient-based consensus on core outcomes guarantees their clinical relevance.^15^ Next, each outcome should be unambiguously defined, preferably according to COSMIN guidelines, and measurement scales identified.^39^ General and disease-specific PROMs already exist to measure HRQoL, such as the Short Form 36 questionnaire and the AneurysmDQoL, respectively.^40,41^ Validated PROMs should similarly be developed for PROs such as “Good communication” and “Adherence to prescribed drugs”.^42^

COS usage can be extended to other areas of implementation, such as mobile health applications for assessment of patients’ health, as in the VASCUL-AID project, and in health registries for quality of care monitoring. With increased developments of data applications and AI in medical practice, concomitantly a need for standardization of outcome measures rises.^43^ COS development enables AI models used in AAA research to align with the framework for fair and trustworthy use of AI (the “FUTURE-AI” guidelines).^44^ Moreover, since a COS will include outcomes that are expert-and patient-approved, its application could be extended for use in (mobile) health applications to ensure reproducibility and clinical relevance of the retrieved health data. To our knowledge, this is the first COS that will be applied in a mobile health application.

A limitation of this study is that the proportion of included postoperative patients exceeded 50% in both the focus groups and the Delphi study (52.6% and 57.3%, respectively). Despite these higher than intended proportions of postoperative patients, the fact that the COS was being developed for small AAAs was emphasized throughout the study. Furthermore, there was limited attendance of patients in the expert consensus meetings, and there was considera-ble attrition (33.1%) across Delphi rounds, both challenges encountered in other COS devel-opments.^11,45^ Attrition was slightly higher for HCPs than for patients (35.9% vs. 28.8%, re-spectively). Response rates are usually >80%, but larger stakeholder groups and higher num-ber of outcomes in the Delphi study are associated with higher rates of attrition.^46^ Future COS development studies may thus benefit from shorter longlists and more personal reminders being sent during the Delphi study.

To conclude, this is the first European COS on small AAAs. These core outcomes should be reported as a minimum in all clinical research on small AAAs. In addition, these outcomes may be applied in cardiovascular risk prediction models for small AAA patients and will be included as endpoints in the VASCUL-AID prediction models.

## Non-standard Abbreviations and Acronyms

AAA: abdominal aortic aneurysm
AI: artificial intelligence
COMET: Core Outcome Measures in Effectiveness Trials
COS: core outcome set
HCP: healthcare professional
HRQoL: health-related quality of life
KOL: key opinion leader
PRO: patient-reported outcome
PROM: patient-reported outcome measure

## Acknowledgements

VASCUL-AID consortium collaborators:

J.M. Wolterink, PhD^1^; I. Išgum, PhD, Prof^2,3,4^; H.A. Marquering, PhD, Prof^3,4^; M.C. Ploem, LL.M, PhD, Prof^5^; F. Catarinella, MD^6^; K.D. Bera MD, PhD^7^; J. Buisan^7^; P. Zhang^7^, J. Raffort, MD, PhD, HDR^8,9,10^; C. Muller, LL.M.^11^; I. Tomic, MD, PhD^12,13^; D. Matejevic, MD^13^; M. Živković, PhD^14^; T. Djuric, PhD^14^; A. Stankovic, PhD^14^; M. Venermo, MD, PhD, Prof^15,16^; M.P. Schijven, MD, PhD, Prof^17,18,19^; B.J.H. van den Born, MD, PhD^20^; R. Delewi, MD, PhD^21,22^; R. Bauersachs, MD, PhD, Prof^23,24^.

1. Department of Applied Mathematics, Technical Medical Centre, University of Twente, Enschede, the Netherlands.

2. Department of Biomedical Engineering and Physics, Amsterdam University Medical Center, Location University of Amsterdam, Amsterdam, The Netherlands.

3. Department of Radiology and Nuclear Medicine, Amsterdam University Medical Center, Location University of Amsterdam, Amsterdam, The Netherlands.

4. Informatics Institute, Faculty of Science, University of Amsterdam, Amsterdam, The Netherlands.

5. Department of Ethics, Law and Humanities, Amsterdam University Medical Center, Location University of Amsterdam, Amsterdam, The Netherlands.

6. Chief Medical Information Officer, Brightfish B.V., Hoofddorp, the Netherlands.

7. Nuffield Department of Surgical Sciences, University of Oxford, Oxford, UK.

8. Clinical Chemistry Laboratory, University Hospital of Nice, France.

9. Institute 3IA Côte d’Azur, Université Côte d’Azur, France.

10. Université Côte d’Azur, CNRS, UMR7370, LP2M, Nice, France.

11. ALLAI, Amsterdam, the Netherlands.

12. Medical Faculty University of Belgrade, Belgrade, Serbia

13. Clinic for Vascular and Endovascular Surgery, Clinical Center of Serbia, Belgrade, Serbia

14. Laboratory for Radiobiology and Molecular Genetics, VINCA Institute of Nuclear Sciences -National Institute of the Republic of Serbia, University of Belgrade, Mike Petrovica Alasa 12-14, P.O. Box 522, Vinca, Belgrade, 11351, Serbia.

15. Department of Vascular Surgery, Helsinki University Hospital, Helsinki, Finland.

16. Department of Vascular Surgery, University of Helsinki, Helsinki, Finland.

17. Amsterdam Public Health, Digital Health, Amsterdam, the Netherlands.

18. Department of Surgery, Amsterdam University Medical Center, Location University of Amsterdam, Amsterdam, the Netherlands.

19. Amsterdam Gastroenterology and Metabolism, Amsterdam, the Netherlands.

20. Department of Public and Occupational Health & Department of Vascular Medicine, Amsterdam Uni-versity Medical Center, Location University of Amsterdam, Amsterdam, The Netherlands.

21. Department of Cardiology, Amsterdam University Medical Center, Location University of Amsterdam, Amsterdam, The Netherlands.

22. Amsterdam Cardiovascular Sciences, Amsterdam, the Netherlands.

23. CCB-Cardioangiologic Center Bethanien, Frankfurt, Germany.

24. Center of Thrombosis and Hemostasis, University of Mainz, Germany.

Expert consensus meeting collaborators

1. P. Aho, MD, PhD^1^; P. Björkman, MD, PhD^2^; M. Lavuori, MD, PhD^2^; J. Fernando Teixeira, MD^3^; C. Martins Barbosa^4^; D. Pereira Dias^4^; C. Dias da Silva^4^; J. Silva Barbosa^4^; M. Markovic, MD, PhD^5^; D. Nio, MD, PhD^6^; M. Warlé, MD, PhD, FEBVS^7^; P. Rygiel, MSc^8^; M. Loncaric, MSc^9,10,11,12^.

1. HUS Abdominal Center, Vascular Surgery and Helsinki University, Helsinki, Finland.

2. Department of Vascular Surgery, HUS Helsinki University Hospital, Helsinki, Finland.

3. Angiology and Vascular Surgery, Unidade de Saúde Local de São João, Porto, Portugal.

4. RISE-Health, Department of Surgery and Physiology, Faculty of Medicine of the University of Porto, Porto, Portugal.

5. Clinic for Vascular and Endovascular Surgery, University Clinical Center of Serbia, Belgrade, Serbia.

6. Department of Vascular Surgery, Spaarne Gasthuis, Haarlem, the Netherlands.

7. Department of Surgery, Division of Vascular and Transplant Surgery, Radboud University Medical Centre, Nijmegen, the Netherlands.

8. Department of Applied Mathematics, Technical Medical Centre, University of Twente, Enschede, The Nether-lands.

9. Department of Biomedical Engineering and Physics, Amsterdam UMC, Meibergdreef 9, Amsterdam, 1105 AZ, The Netherlands

10. Department of Radiology and Nuclear Medicine, Amsterdam UMC, Meibergdreef 9, Amsterdam, 1105 AZ, The Netherlands

11. Informatics Institute, University of Amsterdam, Amsterdam, The Netherlands

12. Amsterdam Cardiovascular Sciences, Amsterdam, The Netherlands

## Disclosures

Kak Khee Yeung, MD, PhD is Editor in Chief of the Journal of Endovascular Therapy.

Riikka Tulamo, MD, PhD is associate editor at European Journal for Vascular & Endovascular Surgery.

## Supplementary materials

Supplementary Tables S1-S5.

